# Pulmonary function and survival one year after dupilumab treatment of acute moderate to severe COVID-19: A follow up study from a Phase IIa trial

**DOI:** 10.1101/2023.09.01.23293947

**Authors:** Jennifer Hendrick, Jennie Z. Ma, Heather M. Haughey, Rachael Coleman, Uma Nayak, Alexandra Kadl, Jeffrey M. Sturek, Patrick Jackson, Mary K. Young, Judith E. Allen, William A. Petri

## Abstract

**Background:** We previously conducted a Phase IIa randomized placebo-controlled trial of 40 subjects to assess the efficacy and safety of dupilumab use in those hospitalized with COVID-19 (NCT04920916). Based on our pre-clinical data suggesting downstream pulmonary dysfunction with COVID-19 induced type 2 inflammation, we contacted patients from our Phase IIa study at 1 year for assessment of Post Covid-19 Conditions (PCC).

**Methods:** Subjects at 1 year after treatment underwent pulmonary function testing (PFTs), high resolution computed tomography (HRCT) imaging, symptom questionnaires, neurocognitive assessments, and serum immune biomarker analysis, with subject survival also monitored. The primary outcome was the proportion of abnormal PFTs, defined as an abnormal diffusion capacity for carbon monoxide (DLCO) or 6-minute walk testing (6MWT) at the 1-year visit.

**Results:** Sixteen of the 29 one-year survivors consented to the follow up visit. We found that subjects who had originally received dupilumab were less likely to have abnormal PFTs compared to those who received placebo (Fisher’s exact p=0.011, adjusted p=0.058). We additionally found that 3 out of 19 subjects (16%) in the dupilumab group died by 1 year compared to 8 out of 21 subjects (38%) in the placebo group (log rank p=0.12). We did not find significant differences in neurocognitive testing, symptoms or CT chest imaging between treatment groups but observed evidence of reduced type 2 inflammation in those who received dupilumab.

**Conclusions:** We observed evidence of reduced long-term morbidity and mortality from COVID-19 with dupilumab treatment during acute hospitalization when added to standard of care regimens.

## Introduction

As COVID-19 infections persist in the United States, they continue to lead to hospitalizations and death due to the increased transmissibility and immune escape characteristics of emerging SARS-CoV-2 variants^1^. Post COVID conditions (PCC), which have led to substantial morbidity and mortality, have been estimated to occur in at least 10% of patients with prior COVID-19 infection with percentages above 50% for those who required hospitalization^2^. Furthermore, repeated COVID-19 infections have been associated with increased death, hospitalization and PCC regardless of vaccination status^3^. Although there are currently no recommended treatment options for PCC, there is evidence that intervention during acute disease can prevent PCC^4^. Further exploration into the immunopathogenesis of this entity is needed to aid the development of additional management strategies.

We have discovered that high plasma interleukin (IL)-13 is associated with COVID-19-induced respiratory failure^5^, a finding further validated in other COVID-19 observational studies^6,7^. IL-13, which signals through the receptor IL-4Rα along with the closely related cytokine IL-4, is involved in eosinophilic inflammation, mucous secretion, goblet cell metaplasia and fibrosis, and has been regularly implicated in atopic disease^8^. We further found that neutralization of IL-13 in K18-hACE2 C57Bl/6J mice protected the animals from severe infection with SARS-CoV-2, as evidenced by reduced clinical score, weight loss and mortality^5^.

RNA-seq analysis of whole lung tissue taken from infected mice who underwent IL-13 neutralization revealed the most downregulated gene to be *Has1*^5^. This gene encodes a synthase responsible for the synthesis of hyaluronan (HA), a major polysaccharide component of the extracellular matrix that has been implicated in viral mediated inflammatory pulmonary diseases, including acute COVID-19^9,10^. This was mechanistically further supported by an increase in HA deposition in human and mouse lung with SARS CoV-2 infection^5^. As HA has been demonstrated as a mediator of multiple pulmonary diseases, including pulmonary fibrosis and idiopathic pulmonary hypertension^11^, this led to the hypothesis that IL-13, acting as a regulator of HA matrix formation, may be involved in pulmonary dysfunction post recovery from COVID-19^12^.

We subsequently conducted a Phase IIa randomized double-blind placebo-controlled trial to assess the safety and efficacy of dupilumab, a monoclonal antibody which blocks IL-4Rα, in mitigating respiratory failure and death in those hospitalized with COVID-19 when added to standard of care regimens^13^. Forty subjects were followed prospectively for 60 days, with collection of clinical outcomes, adverse events, and immunologic biomarkers at multiple time points throughout the study period. Though the primary endpoint of 28-day ventilator-free survival was not reached, we found that subjects randomized to dupilumab had improved 60 day survival as a secondary outcome: 17 out of 19 subjects in the dupilumab group (89%) were alive at day 60 compared to 16 out of 21 in the placebo group (76%)^13^.

Based on our hypothesis that COVID-19-induced type 2 immune activation leads to a downstream destructive pulmonary process, we contacted subjects previously enrolled in our Phase IIa trial, one year after their original enrollment, inviting them to participate in a follow up study for assessment of PCC.

## Methods

### Design

Enrollment in the original Phase IIa trial occurred from June of 2021 until November of 2021 (NCT04920916). Subjects were subsequently contacted to return for follow up visits at 1 year post original enrollment, thus were re-enrolled from August of 2022 until February of 2023. During these visits patients underwent pulmonary function testing (PFTs), high resolution computed tomography (HRCT) imaging, neurocognitive questionnaires, symptom screening and blood sample collection for biomarker analysis. This study was approved by the University of Virginia Institutional Review Board (IRB) in June of 2022.

### Data Collection and Outcomes

PFTs were obtained at the University of Virginia (UVA) Pulmonary Function Testing lab under the supervision of qualified technicians per American Thoracic Society (ATS) guidelines^14^. Testing consisted of spirometry, six-minute walk testing (6MWT) and measurement of diffusion capacity for carbon monoxide (DLCO). All raw PFT data was compared to population-based Global Lung Function Initiative (GLI) predicted values to determine the percentage of predicted value based on age, sex, height, and ethnicity. A PFT measurement at the 5^th^ percentile or lower was considered abnormal^15^.

Neurocognitive testing (Table S1) generated scores were either converted into a T score to allow for comparison to population average and interpreted per instructions in each user manual or interpreted as stated on the questionnaire itself^16–22^. Within the three categories (mood, cognition and functional status/QOL) scores were defined as abnormal if at least one of the tests within the category was deemed a variation from population norm per scoring instructions. Symptom screening included questions for assessment of PCC as listed by the Centers for Disease Control and Prevention (CDC)^23^.

High resolution computed tomography (HRCT) chest scans were scored independently by two pulmonologists with final scores determined by the average of the two. There was negligible interrater variability in scoring that would require a third reviewer. Scores were allotted for visualization of subpleural blebs (score of 0 or 2), bronchiectasis (score of 0 or 2), reticular infiltrates (score from 0-4 depending on extent of parenchymal involvement), ground glass opacities (score from 0-4 depending on extent of parenchymal involvement) and honeycombing (score of 0 or 5) within upper, middle, and lower lung zones. Final scores were determined by the sum of scores from each zone.

Lastly, serum samples were collected from each subject at 1 year for biomarker analysis using a 47-plex cytokine panel (MILLIPLEX SARS-CoV-2 MAP Human Cytokine/Chemokine/Growth Factor Panel). Additionally, serum samples collected at Day 0 from the Phase IIa study were re-analyzed using this panel to eliminate batch effects.

### Statistical Analysis

Data was analyzed with the Chi-square or Fisher’s exact tests for categorical measures (primary outcome, patient characteristics, symptom screening and neurocognitive testing) and two-sample t-test or Wilcoxon rank sum for continuous measures (CT scan scores and serum cytokine, chemokines, and growth factor level differences) to assess for potential differences between groups. Associations between the responses and treatment group were adjusted for pertinent variables via either logistic regression or linear regression analyses while being mindful of cohort sample size.

The primary efficacy endpoint was the proportion of subjects with abnormal PFTs one year after enrollment in Phase IIa study. Abnormal PFTs were defined as either an abnormal DLCO adjusted for hemoglobin level and compared GLI predicted values based on age, sex, height, and ethnicity or an abnormal 6MWT defined as a 3% or more decline in blood oxygen during walking. These PFT components were chosen based on prior ARDS and COVID-19 data^24–26^, which demonstrated these to be the most common post recovery PFT abnormalities found in these patient populations.

A key secondary endpoint was mortality by 1 year which was analyzed as a time to event outcome using Kaplan Meier (KM) survival and Cox regression. For a precision medicine approach, we explored various subgroup analyses by demographics, clinical characteristics, medications received, biomarkers and comorbidities to determine which sub populations would be most likely to benefit from dupilumab. Gender was included in all the Cox regressions due to previously determined imbalance in treatment groups from original study cohort^13^.

As an exploratory endpoint, we created a composite endpoint of either abnormal PFTs at 1 year post enrollment or death in the first year. This outcome was analyzed as a binary endpoint in logistic regression rather than a time-to-event outcome. The decision was made due to the inter-subject variability in follow-up times between those who completed 1-year follow up visits and those who died over the first year. Given this, time from enrollment to PFT measurement was further included in a logistic regression model to control for potential influence of the imbalanced follow-up time.

## Results

Of the 40 originally enrolled patients, there were a total of 11 deaths in the first year (8 in the placebo group, 3 in the dupilumab). Of the remaining 29 survivors at 1 year, 16 patients consented to re-enrollment (Fig S1). There was no difference in demographics, comorbidities, vaccination status, clinical characteristics or medications received during COVID-19 admission between those subjects who consented vs those who declined follow up (Table S2). Among those 16 subjects who consented to 1-year follow up, there was no difference in these characteristics between the two treatment groups (Table 1).

**Table 1:** Demographics, clinical characteristics, co morbidities and medications received during COVID-19 admission of those patients who followed up at 1 year.

|  | Placebo (n=6) | Dupilumab (n=10) |
| --- | --- | --- |
| Age, median (IQR) | 57 (36) | 56 (17) |
| Sex, n male (%) | 4 (67) | 4 (40) |
| BMI, median (IQR) | 31 (10) | 34 (18) |
| Ethnicity, n Hispanic (%) | 1 (10) | 1 (17) |
| Race, n (%) | White: 4 (67)<br>Black: 2 (33)<br>Other: 0 (0) | White: 8 (80)<br>Black: 1 (10)<br>Other: 1 (10) |
| History of cardiac disease, n (%) | 2 (33) | 1 (10) |
| History of kidney disease, n (%) | 1 (17) | 1 (10) |
| History of pulmonary disease, n (%) | 3 (50) | 5 (50) |
| History of neurocognitive dysfunction or stroke, n (%) | 1 (17) | 0 (0) |
| History of smoking, n (%) | 3 (50) | 2 (20) |
| Time till follow up, median (IQR) (%) | 412 (44) | 426 (64) |
| Vaccinated, n (%) | 3 (50) | 5 (50) |
| N protein (ng/mL), median (IQR) | 419 (1000) | 543 (4520) |
| Days from symptom onset to study drug received, median (IQR) | 8 (5) | 9 (4) |
| Days in Hosp, median (IQR) | 5 (8) | 5 (3) |
| Required supplemental oxygen, n (%) | 6 (100) | 9 (90) |
| Required intensive care unit, n (%) | 3 (50) | 1 (10) |
| Required mechanical ventilation, n (%) | 0 (0) | 0 (0) |
| Received monoclonal Ab, n (%) | 2 (33) | 2 (20) |
| Received steroids, n (%) | 6 (100) | 10 (100) |
| Received remdesivir, n (%) | 5 (83) | 8 (80) |
| Received baricitinib, n (%) | 2 (33) | 0 (0) |

We found that all patients in the placebo group (n=6) had an abnormal DLCO or 6MWT at 1-year follow up compared to only 3 of 10 patients in the dupilumab group (Fisher’s exact p=0.011, and p=0.058 after adjusting for time to follow up, preexisting heart/lung disease and smoking history in logistic regression, Fig 1). Though not reaching to statistical significance, there was a trend towards reduced percent predicted forced expiratory volume 1 (FEV1) and percent predicted DLCO in the placebo group compared to the dupilumab group (Fig 2, Table S3).

**Fig 1:**
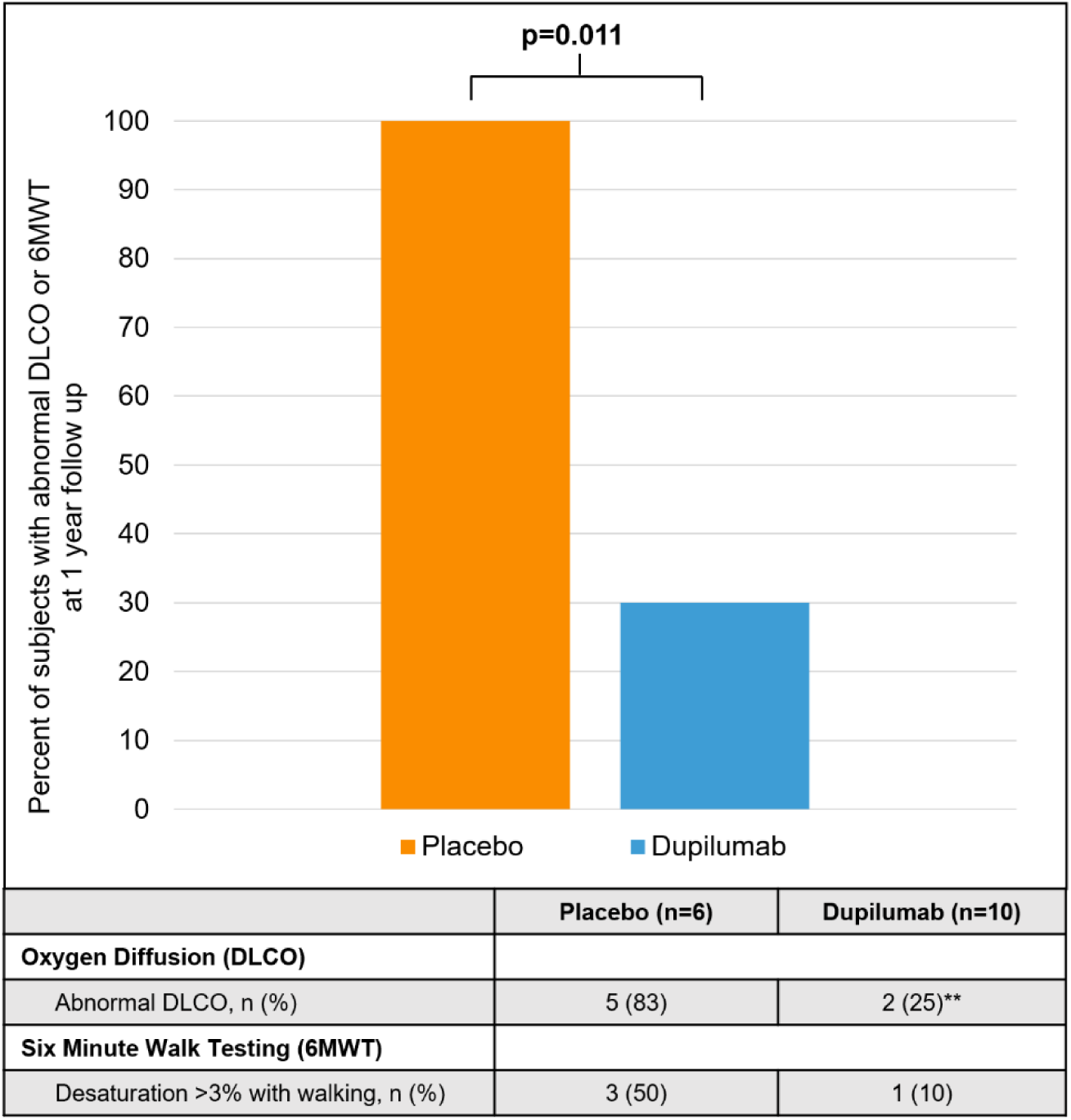
Bar graph illustrating the percent of patients who had abnormal pulmonary function testing (defined by abnormal DLCO or 6MWT) by treatment group at 1-year follow up. The orange box depicts the patients randomized to placebo during initial COVID-19 admission and blue box depicts the subjects randomized to dupilumab during initial COVID-19 admission. Actual number and percentages are displayed in the table at the bottom of the chart. ** indicates two missing values.

**Fig 2:**
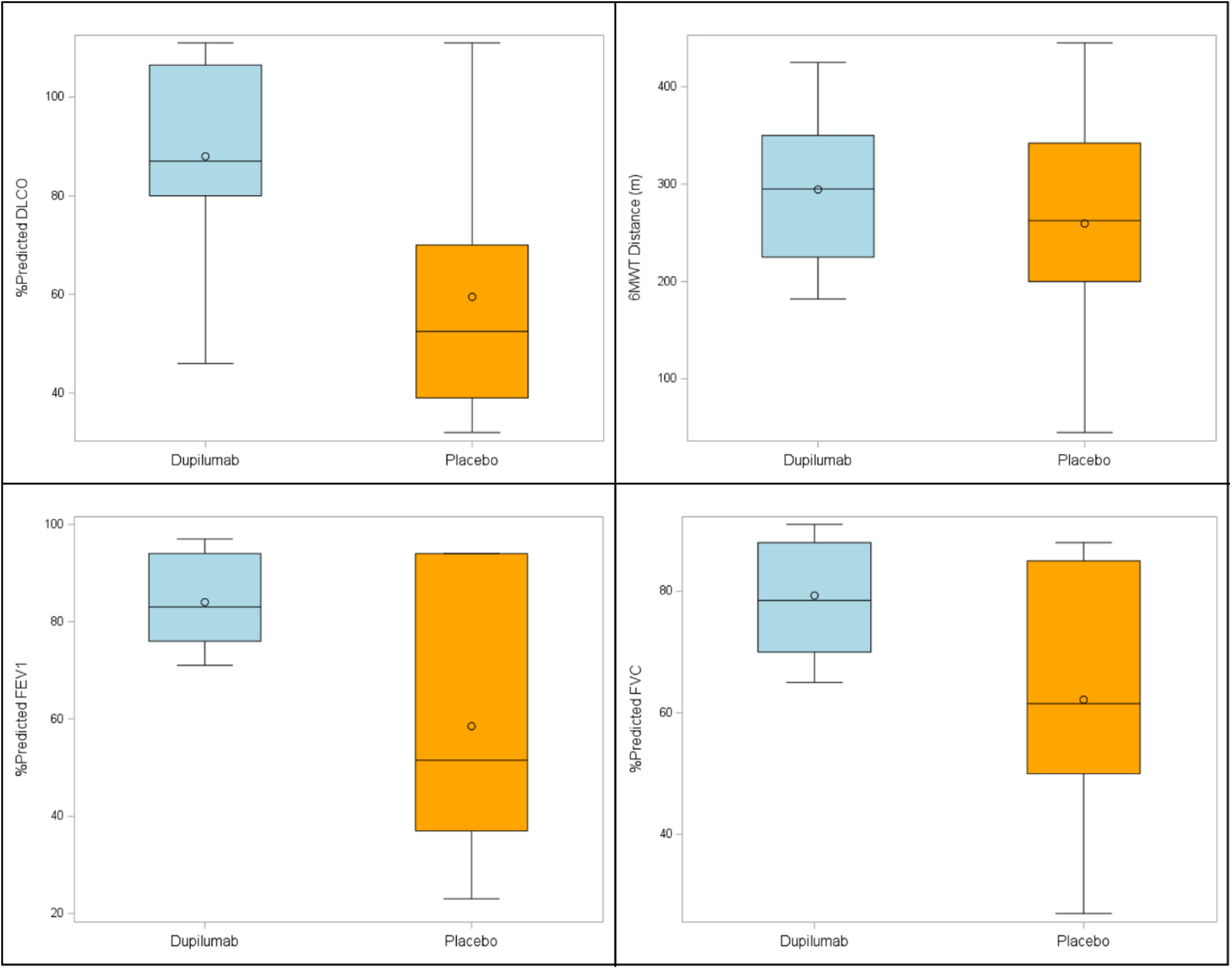
Box plots of percent predicted pulmonary function measures and 6-minute walk test distance (in meters) between treatment groups. The orange boxes depict the patients randomized to placebo and the blue boxes depict the subjects randomized to dupilumab during initial COVID-19 admission. The solid horizontal line within box is representative of median value and the open circle within box is representative of the mean value. FEV1= Forced expiratory volume in 1 second; DLCO= Diffusing capacity of the lungs for carbon monoxide; FVC= Forced vital capacity; 6MWT= Six-minute walk test.

Of the initial 19 subjects randomized to the dupilumab group, 2 died within the first 60 days with 1 additional death occurring thereafter within the 1-year period whereas of the 21 placebo subjects, 5 died in the first 60 days with an additional 3 deaths by 1 year (16% vs 38% total, log rank p=0.12, p=0.25 adjusted for gender via Cox regression, Fig 3). Of the additional 4 deaths that occurred after the Phase IIa study period (60 days), 3 were attributed to respiratory failure (2 occurring during additional hospitalizations and 1 occurring at home) and 1 to septic shock with associated respiratory failure occurring during an additional hospitalization.

**Fig 3:**
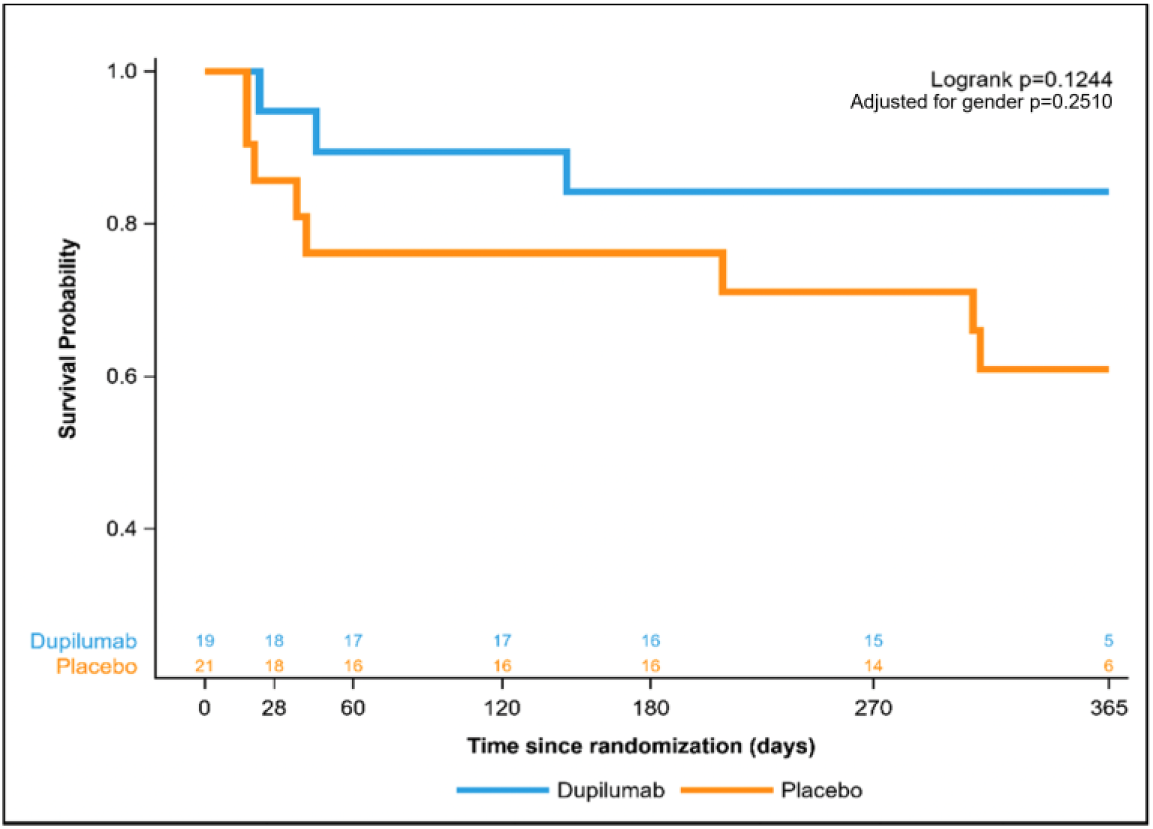
Kaplan Meier curve depicting 1-year mortality between the two treatment groups. Dupilumab group is represented by blue line. Placebo group is represented by the orange line. Adjusted p value indicative of adjustment for sex in the Cox regression.

Through stratification of this analysis by key clinical characteristics we further found that those subjects who had lymphopenia (an established indicator of COVID-19 severity^27^ defined as a lymphocyte count less than 1,000 cells/µL) on the day of original enrollment appeared to respond better to dupilumab compared to placebo in reducing 1-year mortality (log rank p=0.019, and p=0.077 after adjustment for gender via Cox regression, Fig 4).

**Fig 4:**
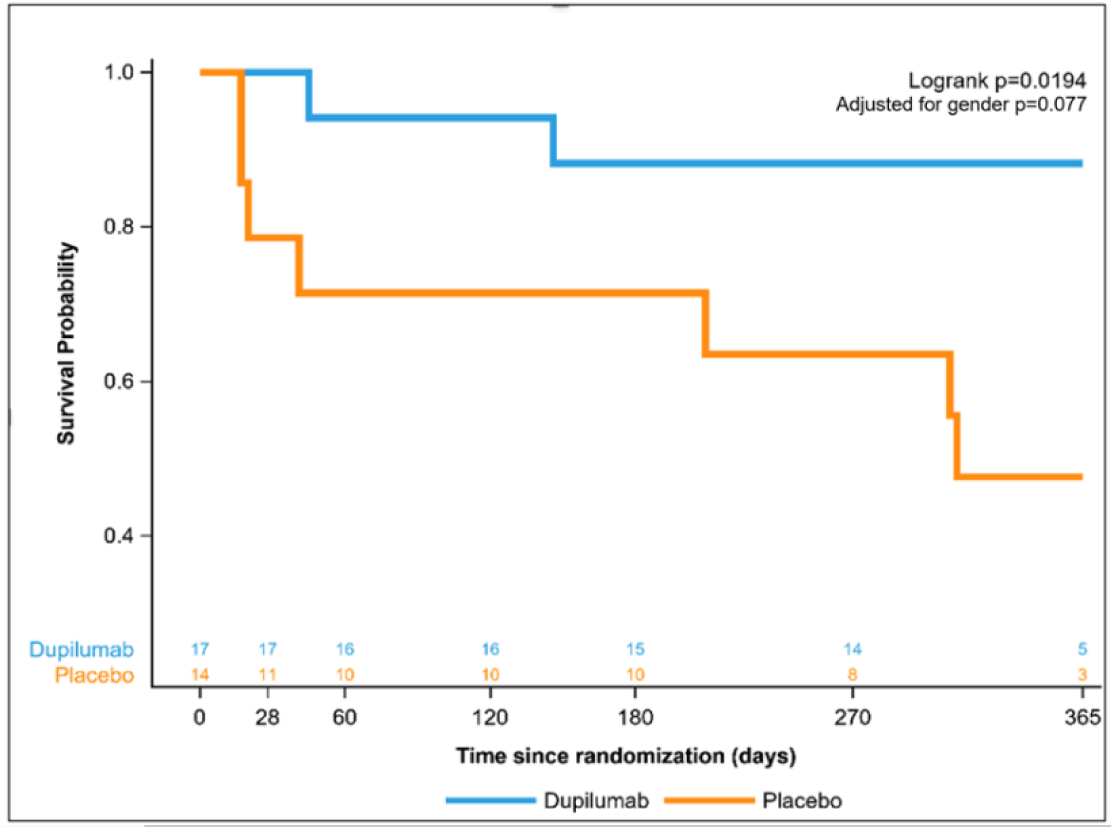
Kaplan Meier curve depicting 1-year mortality between the two treatment groups stratified by lymphopenia (defined as a lymphocyte count less than 1,000 cells/µL). Dupilumab group is represented by blue line. Placebo group is represented by the orange line. Adjusted p value indicative of adjustment for sex in the Cox regression.

For the composite outcome, those who received dupilumab were 77% less likely to die or have abnormal PFTs at 1-year follow up compared to those who received placebo (OR=0.23, 95% CI: 0.06-0.87, p=0.03; and adjusted OR=0.22, p=0.06 after adjusting for age, sex and preexisting pulmonary disease via logistic regression, Fig 5).

**Fig 5:**
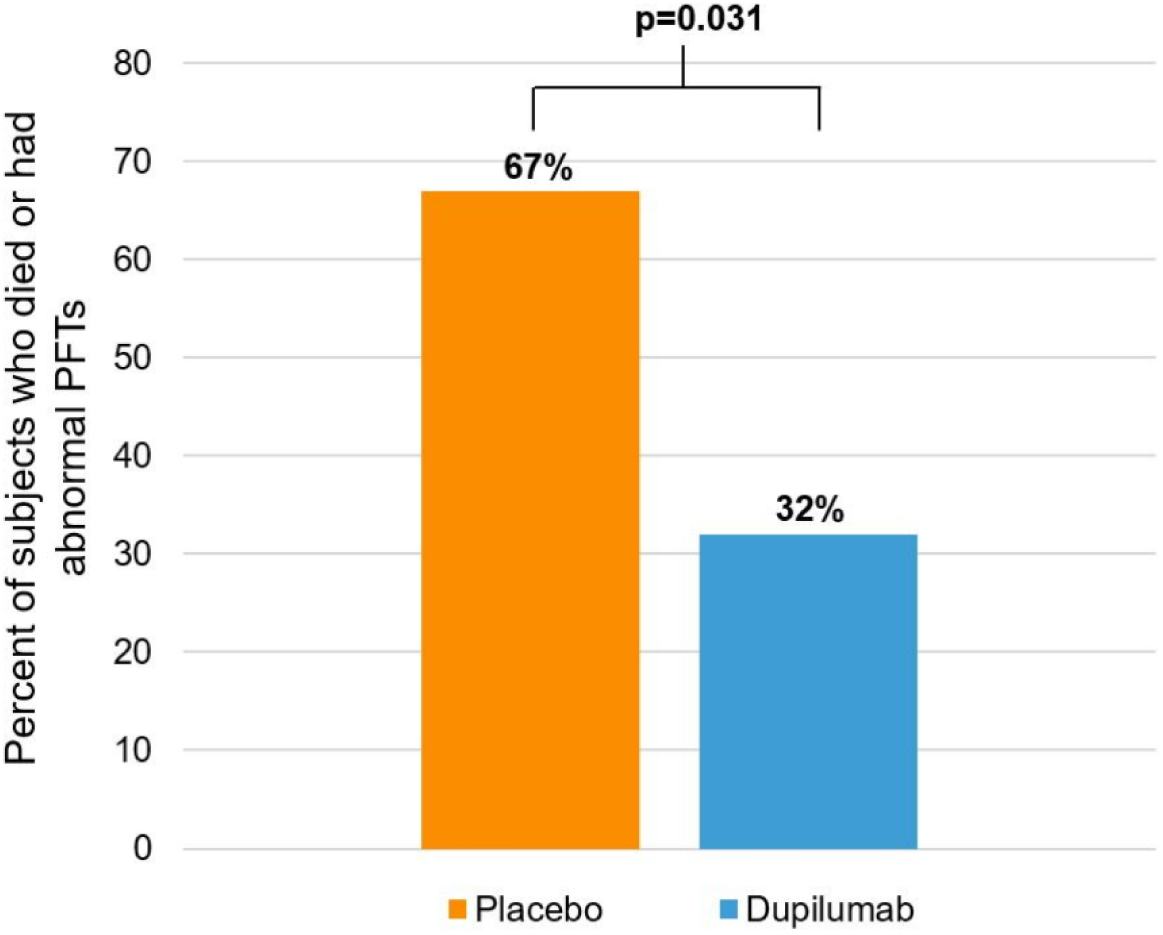
Bar graph illustrating the percent of patients who died or had abnormal pulmonary function testing (defined by abnormal DLCO or 6MWT) by treatment group at 1-year follow up. The orange box depicts the patients randomized to placebo during initial COVID-19 admission and blue box depicts the subjects randomized to dupilumab during initial COVID-19 admission. Actual percent displayed at the top of each bar for corresponding treatment group.

There was no significant difference in proportion of abnormal mood, cognition or functional capacity as determined by neurocognitive testing at 1-year follow up between the two treatment groups (Table S4). When analysis was adjusted for preexisting neurocognitive dysfunction (for analysis of cognition and functional capacity) or preexisting mood disorder (for analysis of mood variable), age (for analysis of cognition and functional capacity variable) and time until follow up appointment (for analysis of all three variables), there remained no significant difference between the groups. Additionally, there was no difference between treatment groups regarding constitutional, mood, respiratory/cardiac, allergic or sensory symptoms at 1-year follow up (Fig S2), which remained without difference when adjusted for pertinent preexisting conditions and time to follow up. HRCT scan scores determined that there was no difference in parenchymal abnormalities between the two treatment groups (Wilcoxon p=0.98, adjusted p=0.96 for preexisting pulmonary disease, smoking status and follow up time via linear regression, Fig S3).

Multiplex analysis of serum cytokines, chemokines and growth factors revealed that those who had normal PFTs at follow up had a larger reduction in eotaxin, tumor necrosis factor (TNF)α and interferon-γ inducible protein (IP)-10 over the year period compared to those with abnormal PFTs. Those with abnormal PFTs also had a larger increase in macrophage inflammatory protein (MIP) 1β at 1-year follow up. Subjects who received dupilumab had a larger decline in eotaxin, interleukin (IL)-12 p40, IP-10, FMS-like tyrosine kinase 3 (FLT3) and a larger increase in IL-1 receptor antagonist (IL-1Ra) over the 1-year period compared to those who received placebo (Fig 6, Table S5).

**Fig 6:**
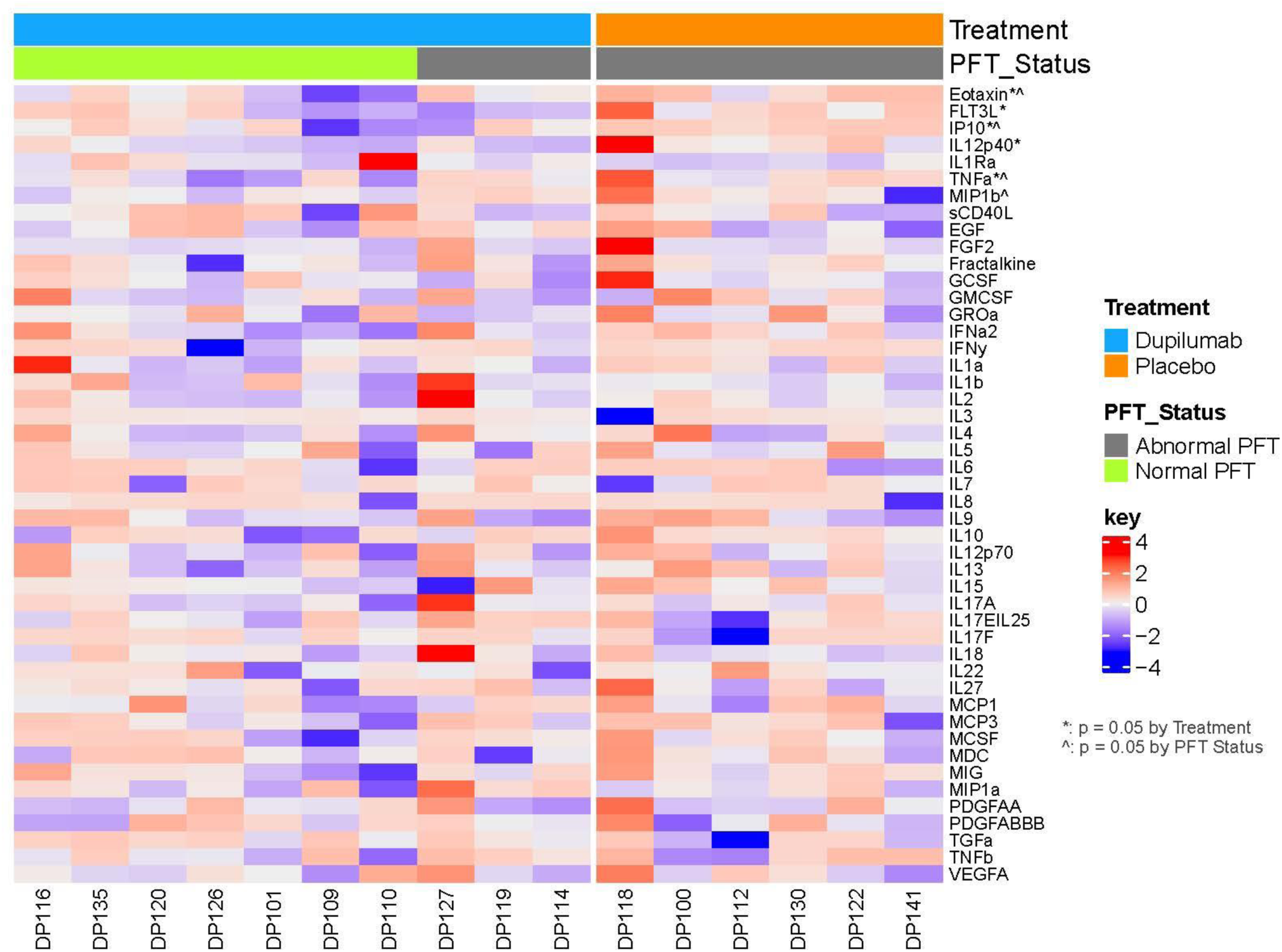
Heatmap showing the standardized differences between baseline and 1-year cytokine, chemokine, or growth factor levels between treatment groups. Each column represents an individual subject. Treatment group is indicated by the top horizontal bar with the blue bar representing the dupilumab group and the orange bar representing the placebo group. Pulmonary function testing (PFTs) status is represented by the second horizontal bar with subjects who had abnormal PFTs at 1 year (defined as abnormal DLCO or 6MWT) represented by the grey bar and those who had normal testing represented by the green bar. Each biomarker is listed on the right of the plot. Those that had significant differences in 1-year changes between treatment groups are marked with a “*” and those with significant differences between 1-year PFT status are marked with a “^”. Note: IL1Ra p=0.065.

## Discussion

In this study we found that those who received IL-13 and IL-4 signaling blockade during their hospitalization for acute COVID-19 infection were less likely to have abnormal pulmonary function at 1 year compared to those who received placebo. Furthermore, as a key secondary outcome, those who initially received dupilumab trended toward reduced 1-year mortality compared to placebo. When stratified by those patients with baseline lymphopenia upon initial trial enrollment, there was a statistically significant difference in 1-year mortality between treatment groups. All of these findings suggest a long-term benefit to IL-4Rα blockade during acute, severe COVID-19 infection.

Treatment with dupilumab for acute COVID-19 was also associated with lower biomarkers for both type 1 and type 2 inflammation, with an increase in the anti-inflammatory cytokine IL-1Ra. Previous studies have demonstrated persistent activation of T cells and innate immune cells for months post COVID-19 recovery, demonstrating prolonged immune activation in those with PCC^28,29^. Although this might suggest the need for prolonged anti-inflammatory therapeutics for prevention of PCC, our study suggests that an acute type 2 immune blockade may deter a prolonged and destructive downstream immune cascade.

Type 2 inflammation has been established as a significant contributor to pathogenesis in subsets of asthma and chronic obstructive pulmonary disease (COPD) patients with eosinophilia, and dupilumab has been demonstrated to be a beneficial therapeutic in both entities^30,31^. As those studies involved multiple treatments during the chronic stage of disease, what is striking here is that short-term treatment during acute COVID-19 appeared to have longer term benefits.

Although no subjects originally enrolled in the trial presented with peripheral eosinophilia, when stratified by baseline lymphopenia, we found a significantly reduced mortality in the dupilumab group compared to placebo. While the mechanism of lymphopenia, whether it is due to viral induced T cell apoptosis or increased tissue uptake of lymphocytes, is unknown, it has been associated with increased inflammation and worse outcomes in COVID-19 patients^27,32^. This suggests that dupilumab may be most beneficial for those in the significant COVID-19 induced inflammatory state that leads to hospitalization.

Specific PFT abnormality differences related to oxygen diffusion seen in our study suggests downstream alterations at the alveolar-capillary interface, which as hypothesized in the literature, could be possibly explained by persistent endothelial activation and fibrosis^33^. We saw a non-significant trend towards reduced FEV1 in our dupilumab group compared to placebo. Recognizing the power limitations in our study, we would have expected FEV1 to be the predominant PFT difference seen in this study based on prior dupilumab studies in COPD and asthma^31,34^. Our findings suggest that dupilumab was primarily able to inhibit a downstream vascular or perivascular fibrotic process induced by severe COVID-19, perhaps as a consequence of altered HA deposition^5^.

We found evidence of diminished type 2 immune activity in our treatment group compared to placebo at 1 year through observation of larger changes in peripheral eotaxin levels, a chemokine which functions in the recruitment of eosinophils to promote eosinophil induced inflammation^35^. We further found larger eotaxin changes in those who had normal PFTs at 1-year follow up compared to those with abnormal PFTs, further supporting the association between diminished type 2 immune activity and normal pulmonary function post COVID-19. This trend was also visualized with IP-10, a chemokine downstream of interferon gamma (IFNγ)^36^, and TNF-α, a cytokine implicated in chronic inflammation, autoimmunity and also associated with PCC^33,37^. This further emphasizes dupilumab’s anti-inflammatory effect and potential association with improved pulmonary function post COVID-19.

PFTs in our 1-year study cohort were similar to Wu et al. who observed that a third of patients hospitalized without mechanical ventilation for their COVID-19 illness exhibited reduced DLCO at 1-year follow up^26^. Eberst et al. only found 11% of patients with abnormal DLCO and 38% of abnormal 6MWT at 1-year follow up in their cohort of patients admitted to the ICU for their COVID-19 illness^25^. Comparison to our cohort (44% abnormal DLCO, 25% abnormal 6MWT in the cohort as a whole) suggests modestly higher rates of pulmonary dysfunction in our study population. This is consistent with our Phase IIa findings, where we noted a higher disease severity in our subjects compared to other reports during the delta wave^13^. Differences in our PFT findings from other studies may be due to the variation pulmonary function reference ranges used, PFT lab variability, SARS-CoV-2 variants involved (both observational studies included cohorts enrolled pre delta wave), pre-existing comorbidities of subjects or differences in COVID-19 management at the respective places and times. The 1-year mortality in our study was comparable to others as we found a 38% 1-year mortality in our placebo group compared to 30-50% seen in other studies assessing those hospitalized for COVID-19^38,39^.

Interestingly we did not see a difference in reported symptoms, neurocognitive assessments, and CT findings between the two treatment groups as we might have expected given the differences found in PFTs. Inconsistencies between subject perception of post COVID-19 symptoms and measured pulmonary function measures has also been demonstrated in other post COVID-19 studies^25,33,40^. As our study subjects did not have pre COVID assessments of these parameters for comparison, a possible explanation may be due to our inability to capture relative change in symptoms and PFTs from baseline measures. As we defined pulmonary function abnormalities based on population-based norms, it is possible that patients suffered relative functional decline in their measures which did not reach our pre-defined abnormality thresholds. That said, these subthreshold changes would likely require further study to determine their clinical implications. Lack of differences seen between CT scans may be reflective of vascular or microscopic parenchymal abnormalities that are not detectable on pulmonary imaging.

Limitations to this study include the small sample size generated by the significant mortality in our placebo group in addition to the subjects who declined follow up visits. Regarding the latter, there were no differences that would suggest differences in clinical severity (either limiting functional capability to attend appointments or interest in follow up appointments due to lack of clinical symptoms) or demographics (age, sex, ethnicity, race) between those who followed up compared to those who declined.

Acknowledging these limitations, the extent of separation between treatment groups seen in this study underscores the need for a larger, multicenter trial for further validation. Further supporting the need for additional study are the extensive pre-clinical human and mouse data demonstrating the significant contribution of a type 2 immune response to COVID-19 induced respiratory dysfunction, the findings from our Phase IIa randomized control trial, the favorable safety profile of dupilumab and the therapeutic efficacy of dupilumab when used in other inflammatory pulmonary conditions. We conclude that dupilumab has promise as a therapeutic option in preventing post-COVID morbidity and mortality.

## Data Availability

All data produced in the present study are available upon reasonable request to the authors.

## ACKNOWLEDGEMENTS

We wish to thank the patients who consented to enroll in this study to help others with post-COVID-19 conditions and the staff at the University of Virginia Post COVID clinic for their assistance with this follow up study. We thank Amy Warren and Lori Elder for IRB protocol preparation and IND preparation, submission and maintenance. We would also like to thank the Biorepository and Tissue Research Facility at UVA who collected, organized, and analyzed research samples.

## Works Cited

1. Ma KC, Dorabawila V, León TM, et al. Trends in Laboratory-Confirmed SARS-CoV-2 Reinfections and Associated Hospitalizations and Deaths Among Adults Aged ≥18 Years — 18 U.S. Jurisdictions, September 2021–December 2022. MMWR Morb Mortal Wkly Rep. 2023;72(25):683–689. doi:10.15585/mmwr.mm7225a3

2. Bull-Otterson L, Baca S, Saydah S, et al. Post–COVID Conditions Among Adult COVID-19 Survivors Aged 18–64 and ≥65 Years — United States, March 2020–November 2021. MMWR Morb Mortal Wkly Rep. 2022;71(21):713–717. doi:10.15585/mmwr.mm7121e1

3. Bowe B, Xie Y, Al-Aly Z. Acute and postacute sequelae associated with SARS-CoV-2 reinfection. Nat Med. 2022;28(11):2398–2405. doi:10.1038/s41591-022-02051-3

4. Bramante CT, Buse JB, Liebovitz DM, et al. Outpatient treatment of COVID-19 and incidence of post-COVID-19 condition over 10 months (COVID-OUT): a multicentre, randomised, quadruple-blind, parallel-group, phase 3 trial. Lancet Infect Dis. Published online June 2023. doi:10.1016/S1473-3099(23)00299-2

5. Donlan AN, Sutherland TE, Marie C, et al. IL-13 is a driver of COVID-19 severity. JCI Insight. 2021;6(15). doi:10.1172/jci.insight.150107

6. Liang Y, Ge Y, Sun J. IL-33 in COVID-19: friend or foe? Cell Mol Immunol. 2021;18(6):1602–1604. doi:10.1038/s41423-021-00685-w

7. Lucas C, Wong P, Klein J, et al. Longitudinal analyses reveal immunological misfiring in severe COVID-19. Nature. 2020;584(7821):463–469. doi:10.1038/s41586-020-2588-y

8. Wynn TA. IL-13 Effector Functions. Annu Rev Immunol. 2003;21(1):425–456. doi:10.1146/annurev.immunol.21.120601.141142

9. Bell TJ, Brand OJ, Morgan DJ, et al. Defective lung function following influenza virus is due to prolonged, reversible hyaluronan synthesis. Matrix Biol. 2019;80:14–28. doi:10.1016/j.matbio.2018.06.006

10. Albtoush N, Petrey AC. The role of hyaluronan synthesis and degradation in the critical respiratory illness COVID-19. Am J Physiol Physiol. 2022;322(6):C1037–C1046. doi:10.1152/ajpcell.00071.2022

11. Lennon FE, Singleton PA. Role of hyaluronan and hyaluronan-binding proteins in lung pathobiology. Am J Physiol - Lung Cell Mol Physiol. 2011;301(2). doi:10.1152/ajplung.00071.2010

12. Sasson J, Moreau GB, Petri WA. The role of interleukin 13 and the type 2 immune pathway in COVID-19. *Ann Allergy*, Asthma Immunol. Published online March 2023. doi:10.1016/j.anai.2023.03.009

13. Sasson J, Donlan AN, Ma JZ, et al. Safety and Efficacy of Dupilumab for the Treatment of Hospitalized Patients With Moderate to Severe Coronavirus Disease 2019: A Phase 2a Trial. Open Forum Infect Dis. 2022;9(8). doi:10.1093/ofid/ofac343

14. Stanojevic S, Kaminsky DA, Miller MR, et al. ERS/ATS technical standard on interpretive strategies for routine lung function tests. Eur Respir J. 2022;60(1):2101499. doi:10.1183/13993003.01499-2021

15. Cooper BG, Stocks J, Hall GL, et al. The Global Lung Function Initiative (GLI) Network: bringing the world’s respiratory reference values together. Breathe. 2017;13(3):e56–e64. doi:10.1183/20734735.012717

16. Julayanont P, Tangwongchai S, Hemrungrojn S, et al. The Montreal Cognitive Assessment-Basic: A Screening Tool for Mild Cognitive Impairment in Illiterate and Low-Educated Elderly Adults. J Am Geriatr Soc. 2015;63(12):2550–2554. doi:10.1111/jgs.13820

17. Morin CM. Insomnia Severity Index (ISI).; 1993.

18. Wallace M, Shelkey M. Katz Index of Independence in Activities of Daily Living (ADL).; 2007.

19. Oemar M, Janssen B. EQ-5D-5L User Guide.; 2013.

20. Cella D, Riley W, Stone A, et al. The Patient-Reported Outcomes Measurement Information System (PROMIS) developed and tested its first wave of adult self-reported health outcome item banks: 2005–2008. J Clin Epidemiol. 2010;63(11):1179–1194. doi:10.1016/j.jclinepi.2010.04.011

21. PROMIS® Scoring Manuals and Score Cut Points. Accessed July 19, 2023. https://www.healthmeasures.net/promis-scoring-manuals

22. User Manual for the Quality of Life in Neurological Disorders (Neuro-QoL) Measures.; 2015.

23. Centers for Disease Control and Prevention. Post-COVID Conditions: Information for Healthcare Providers. https://www.cdc.gov/coronavirus/2019-ncov/hcp/clinical-care/post-covid-conditions.html#assessment-and-testing.

24. Herridge MS, Cheung AM, Tansey CM, et al. One-Year Outcomes in Survivors of the Acute Respiratory Distress Syndrome. N Engl J Med. 2003;348(8):683–693. doi:10.1056/NEJMoa022450

25. Eberst G, Claudé F, Laurent L, et al. Result of one-year, prospective follow-up of intensive care unit survivors after SARS-CoV-2 pneumonia. Ann Intensive Care. 2022;12(1). doi:10.1186/s13613-022-00997-8

26. Wu X, Liu X, Zhou Y, et al. 3-month, 6-month, 9-month, and 12-month respiratory outcomes in patients following COVID-19-related hospitalisation: a prospective study. Lancet Respir Med. 2021;9(7):747–754. doi:10.1016/S2213-2600(21)00174-0

27. Tan L, Wang Q, Zhang D, et al. Lymphopenia predicts disease severity of COVID-19: a descriptive and predictive study. Signal Transduct Target Ther. 2020;5(1):16–18. doi:10.1038/s41392-020-0148-4

28. Phetsouphanh C, Darley DR, Wilson DB, et al. Immunological dysfunction persists for 8 months following initial mild-to-moderate SARS-CoV-2 infection. Nat Immunol. 2022;23(2):210–216. doi:10.1038/s41590-021-01113-x

29. Glynne P, Tahmasebi N, Gant V, Gupta R. Long COVID following Mild SARS-CoV-2 Infection: Characteristic T Cell Alterations and Response to Antihistamines. J Investig Med. 2022;70(1):61–67. doi:10.1136/jim-2021-002051

30. Corren J, Castro M, O’Riordan T, et al. Dupilumab Efficacy in Patients with Uncontrolled, Moderate-to-Severe Allergic Asthma. J Allergy Clin Immunol Pract. 2020;8(2):516–526. doi:10.1016/j.jaip.2019.08.050

31. Bhatt SP, Rabe KF, Hanania NA, et al. Dupilumab for COPD with Type 2 Inflammation Indicated by Eosinophil Counts. N Engl J Med. 2023;389(3):205–214. doi:10.1056/NEJMoa2303951

32. Wang X, Liu Z, Lu L, Jiang S. The putative mechanism of lymphopenia in COVID-19 patients. J Mol Cell Biol. 2022;14(5). doi:10.1093/jmcb/mjac034

33. Altmann DM, Whettlock EM, Liu S, Arachchillage DJ, Boyton RJ. The immunology of long COVID. Nat Rev Immunol. Published online July 11, 2023. doi:10.1038/s41577-023-00904-7

34. Wenzel S, Castro M, Corren J, et al. Dupilumab efficacy and safety in adults with uncontrolled persistent asthma despite use of medium-to-high-dose inhaled corticosteroids plus a long-acting β2 agonist: a randomised double-blind placebo-controlled pivotal phase 2b dose-ranging trial. Lancet. 2016;388(10039):31–44. doi:10.1016/S0140-6736(16)30307-5

35. Conroy DM, Williams TJ. Eotaxin and the attraction of eosinophils to the asthmatic lung. Respir Res. 2001;2(3):150–156. doi:10.1186/rr52

36. Liu M, Guo S, Hibbert JM, et al. CXCL10/IP-10 in infectious diseases pathogenesis and potential therapeutic implications. Cytokine Growth Factor Rev. Published online July 2011. doi:10.1016/j.cytogfr.2011.06.001

37. van Loo G, Bertrand MJM. Death by TNF: a road to inflammation. Nat Rev Immunol. 2023;23(5):289–303. doi:10.1038/s41577-022-00792-3

38. Mainous AG, Rooks BJ, Wu V, Orlando FA. COVID-19 Post-acute Sequelae Among Adults: 12 Month Mortality Risk. Front Med. 2021;8. doi:10.3389/fmed.2021.778434

39. Novelli L, Raimondi F, Carioli G, et al. One-year mortality in COVID-19 is associated with patients’ comorbidities rather than pneumonia severity. Respir Med Res. 2023;83. doi:10.1016/j.resmer.2022.100976

40. Lehmann A, Gysan M, Bernitzky D, et al. Comparison of pulmonary function test, diffusion capacity, blood gas analysis and CT scan in patients with and without persistent respiratory symptoms following COVID-19. BMC Pulm Med. 2022;22(1):196. doi:10.1186/s12890-022-01987-z

