## Supplemental Data for "Pulmonary function and survival one year after dupilumab treatment of acute moderate to severe COVID-19: A follow up study from a Phase IIa trial"

| **Category** | **Test Name** |
| --- | --- |
| Mood | - Patient-Reported Outcomes Measurement Information System (PROMIS) Anxiety - PROMIS Depression |
| Cognition | - Montreal Cognitive Assessment (MOCA) - Neuro- Quality of Life (QoL) |
| Functional Status/ Quality of Life | - Insomnia Severity Index (ISI) - Katz Index of Independence In Activities of Daily Living (Katz-ADL) - EuroQOL (EQ)-5D-5L |

Table S1: Neurocognitive tests administered at 1 year follow up visits.

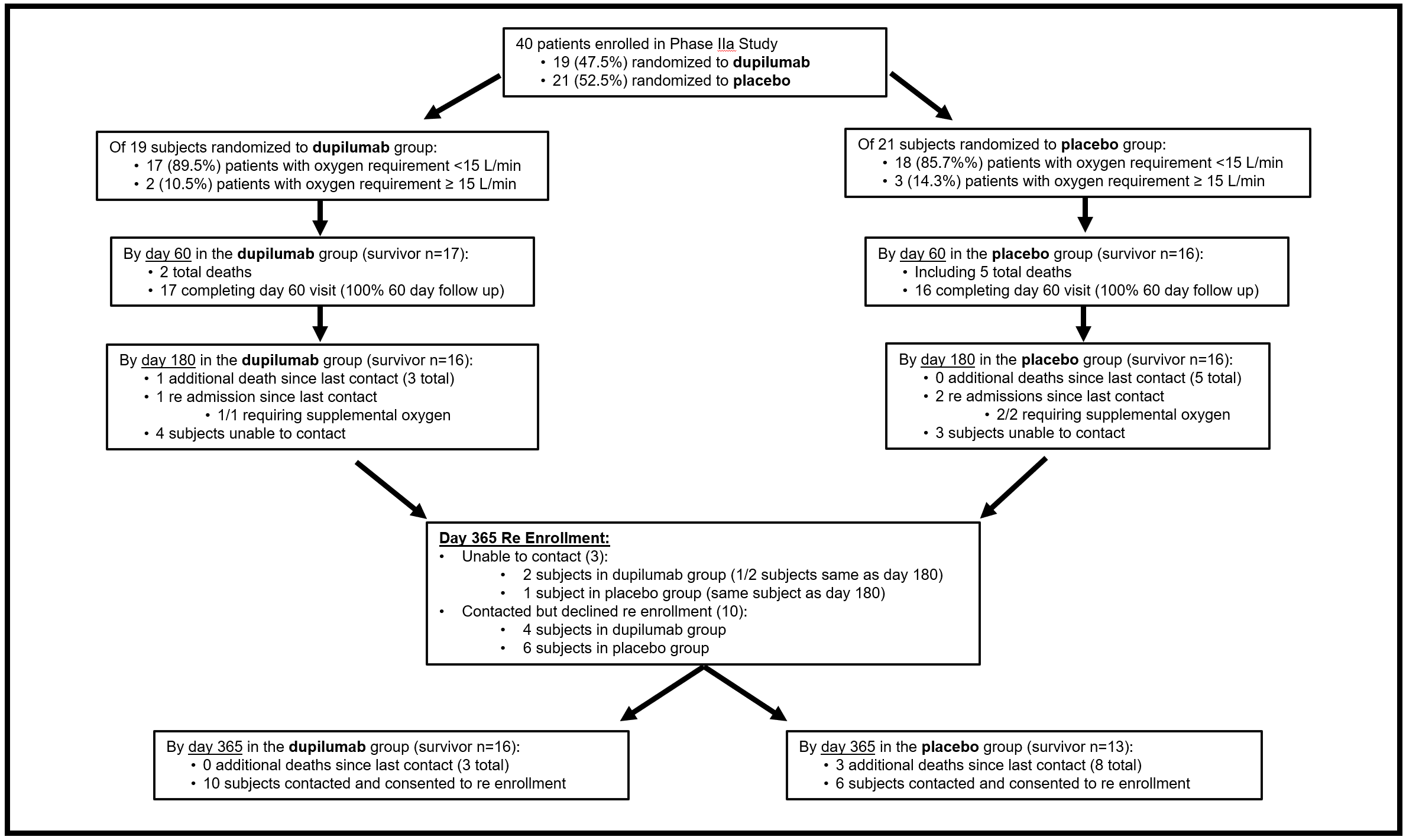

Fig S1: Study timeline and re-enrollment.

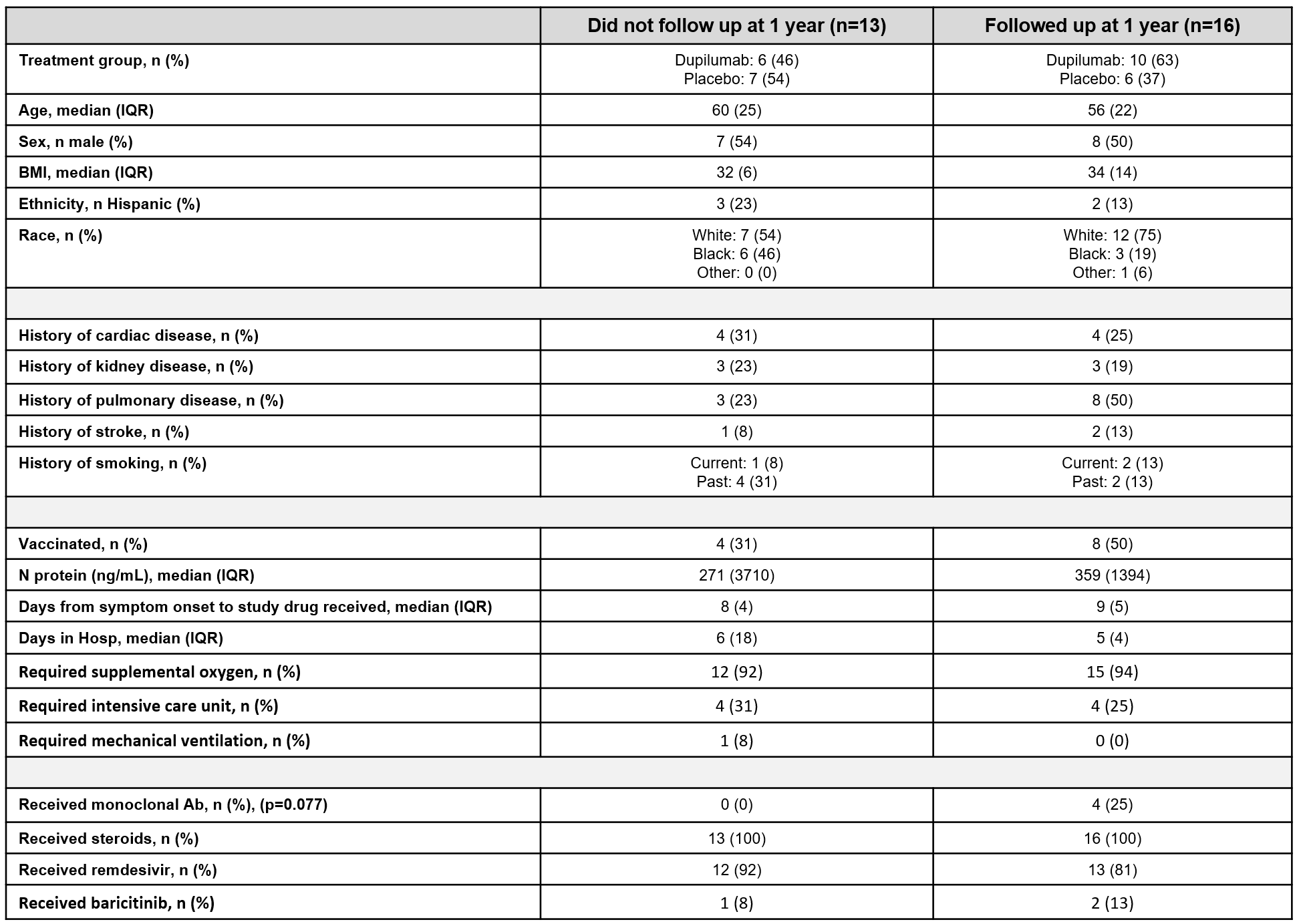

Table S2: Demographics, co morbidities, clinical characteristics and medications received while during COVID-19 admission between those who consented to follow up visit versus those who were alive and declined follow up visit at 1 year post enrollment.

Table S3: Pulmonary function testing measurements by treatment group at 1 year follow up. Spirometry and oxygen diffusion data is displayed as a continuous measure indicating percent of predicted compared to Global Lung Function Initiative (GLI) predicted values based on age, sex, height and ethnicity. This data is also displayed as a binary measure (% abnormal) which depicts the interpretation of measurements. Six-minute walk testing is displayed as continuous measures for actual distance walked and percent of predicted as determined by subject age, sex, height and weight. It is also displayed as a binary variable indicating percent of subjects who desaturated, who reported dyspnea and who displayed exercise limitation during their six-minute walk. * indicates 1 missing value ** indicates 2 missing values.

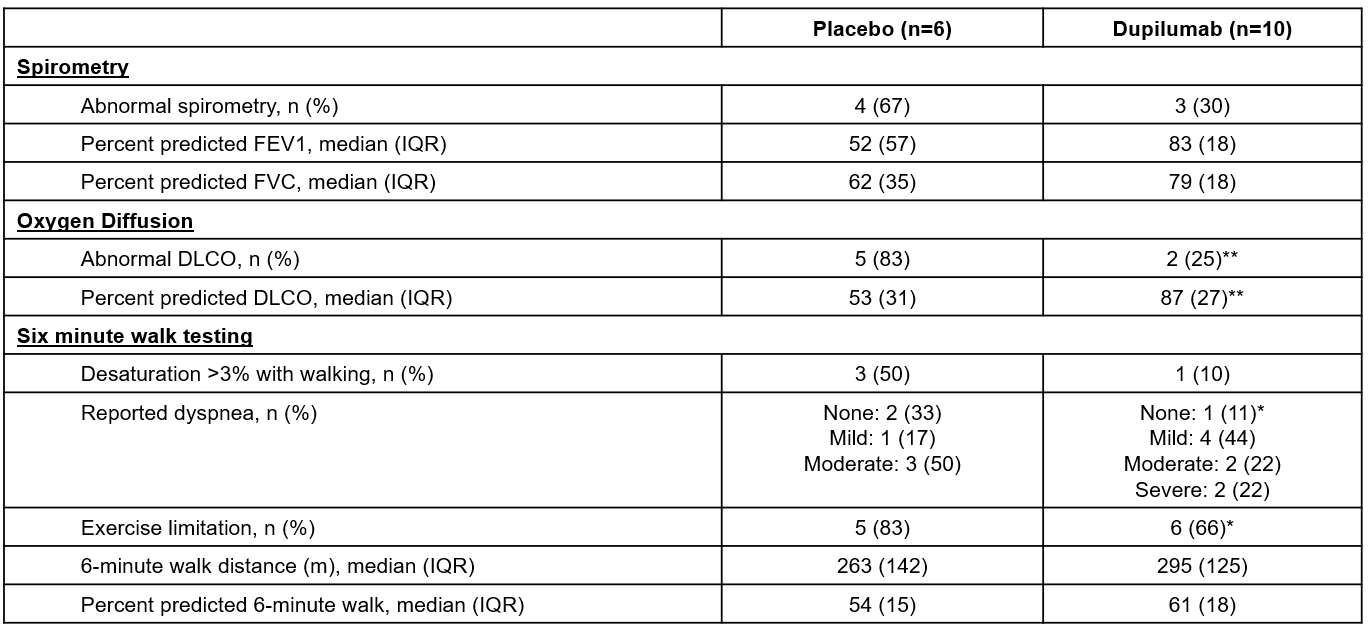

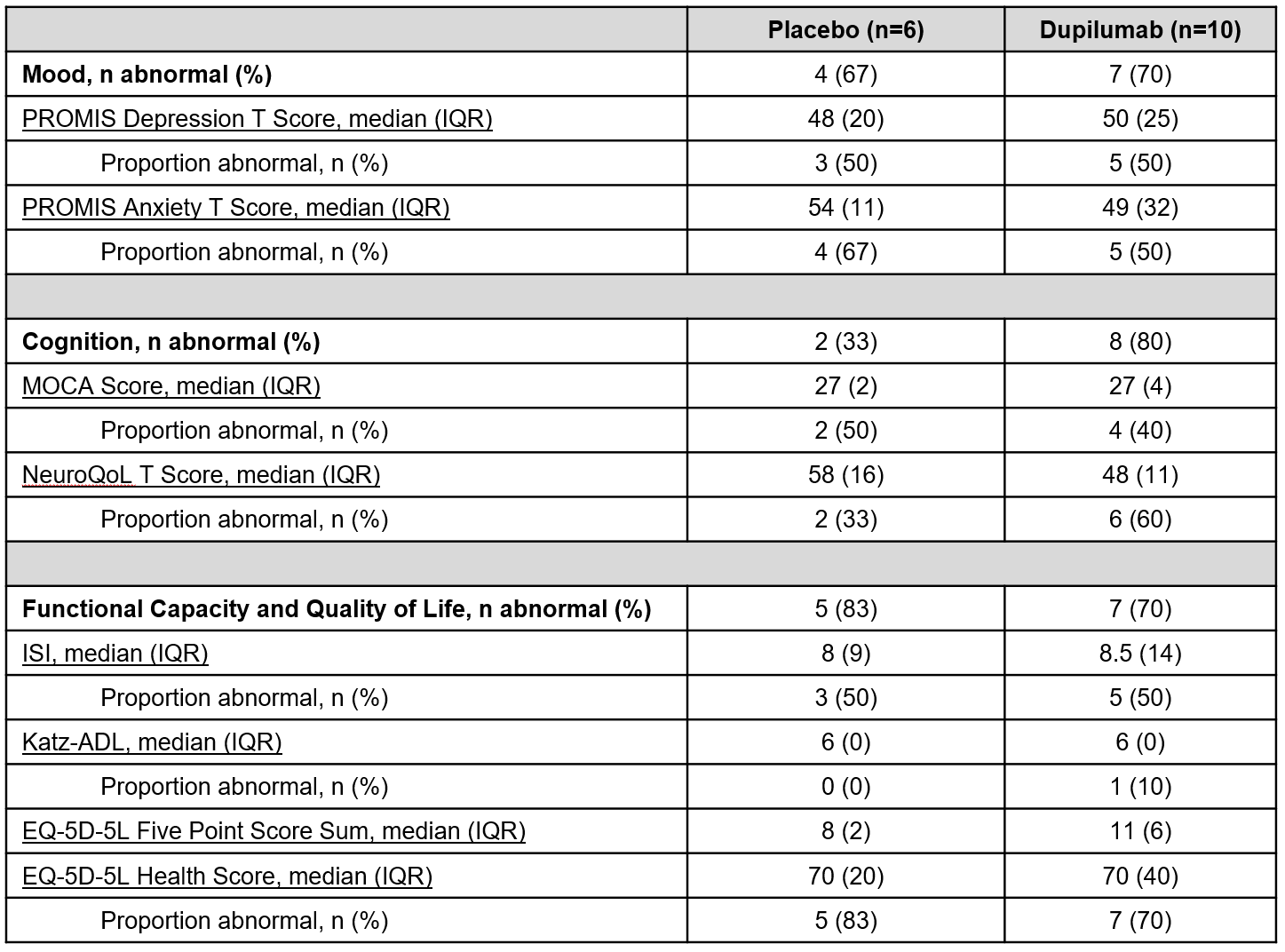

Table S4: Neurocognitive testing results at 1 year follow up by treatment group. Each test is split into three types of measurement categories: mood, cognition and functional capacity. Measurements are displayed as a score and interpretation (% abnormal) of those scores for that treatment group. The EQ-5D-5L five-point score raw numbers are displayed as a median of the sum of all 5 scores and as a median of reported health score (on scale from 0-100) for each treatment group.

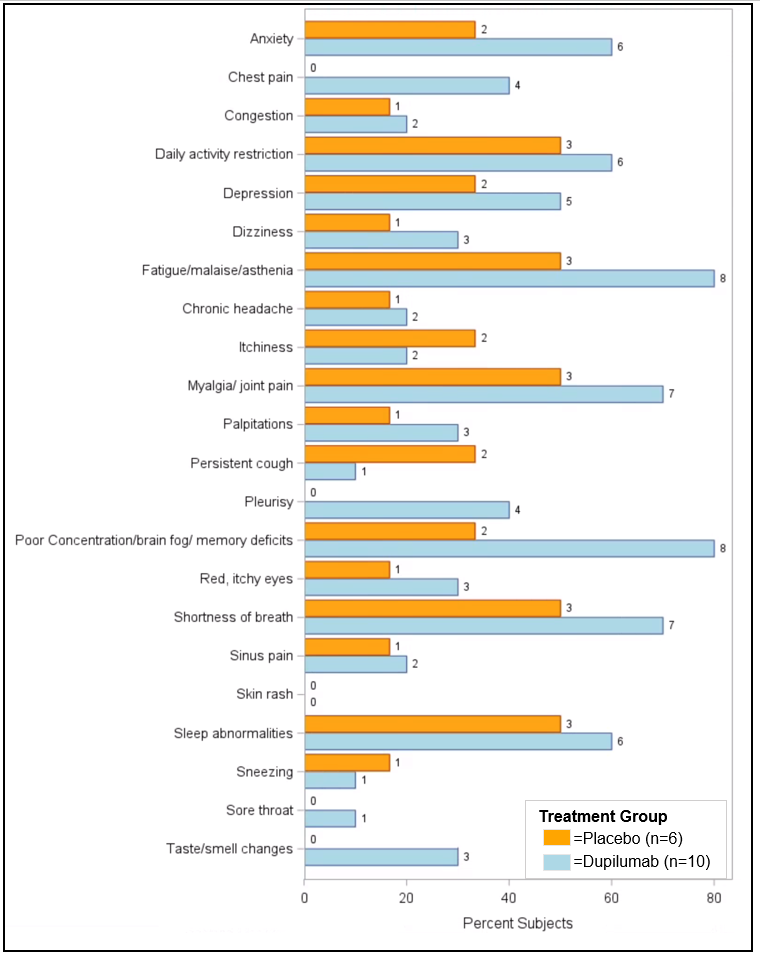

Fig S2: Symptoms reported at 1 year follow up by treatment group. The horizontal axis indicates the percent of patients who followed up and reported that symptom. The number at the end of each bar represents raw count out of the total for that treatment group. The orange boxes depict the patients randomized to placebo during initial COVID-19 admission and blue box depicts the subjects randomized to dupilumab during initial COVID-19 admission.

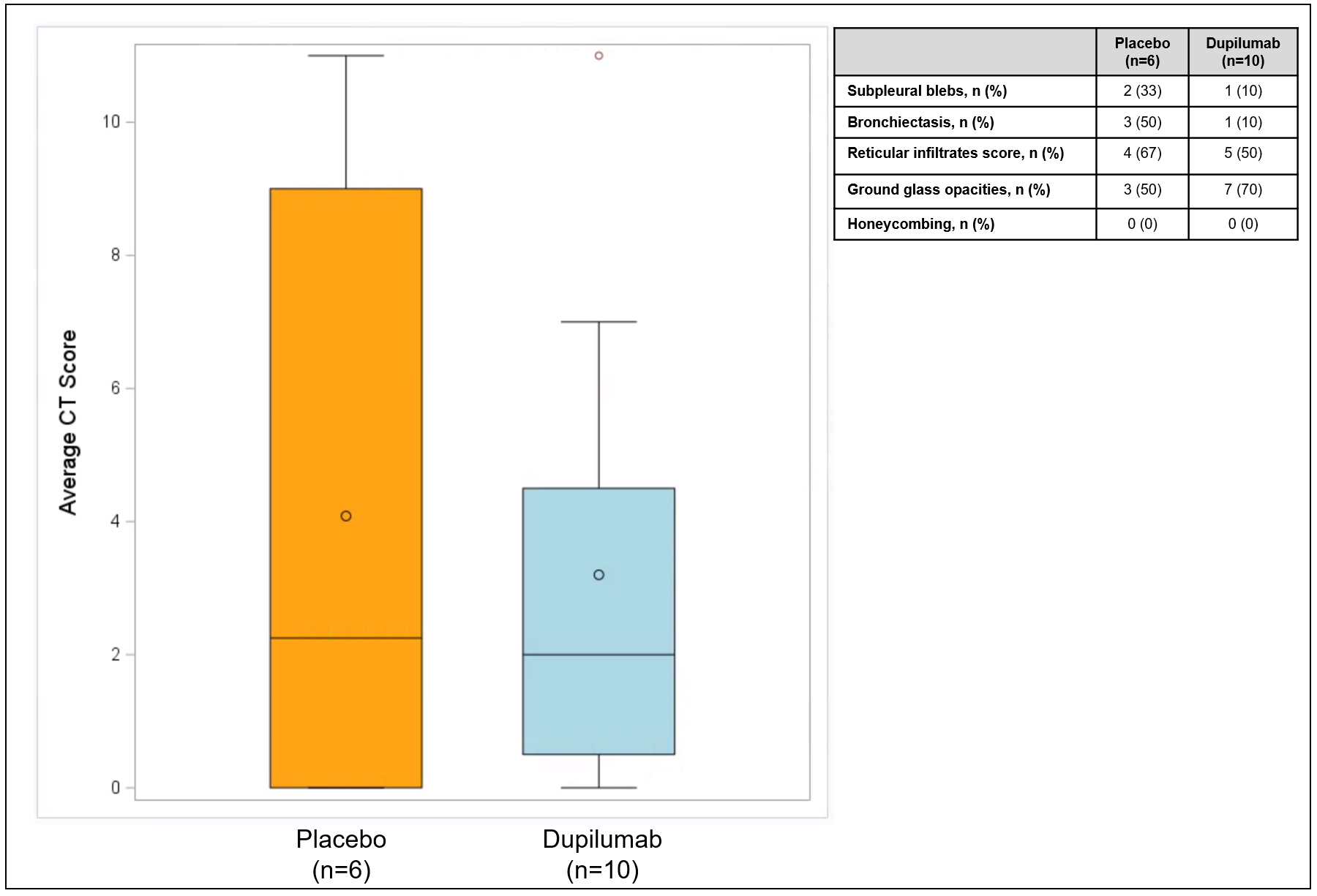

Fig S3: Computer tomography (CT) scores and abnormalities seen by treatment group. CT scores are depicted in the box and whisker plot on the left and were generated as an average of scores from two independent pulmonologists. Scores were generated by totaling scores based on parenchymal findings from three areas of the lung. The orange box depicts the patients randomized to placebo and the blue box depicts the subjects randomized to dupilumab during initial COVID-19 admission. Solid horizontal line within box is representative of median value and open circle within box is representative of mean value. The table on the right shows number (n) and percentage (%) of subjects who displayed those parenchymal abnormalities at any lung level.

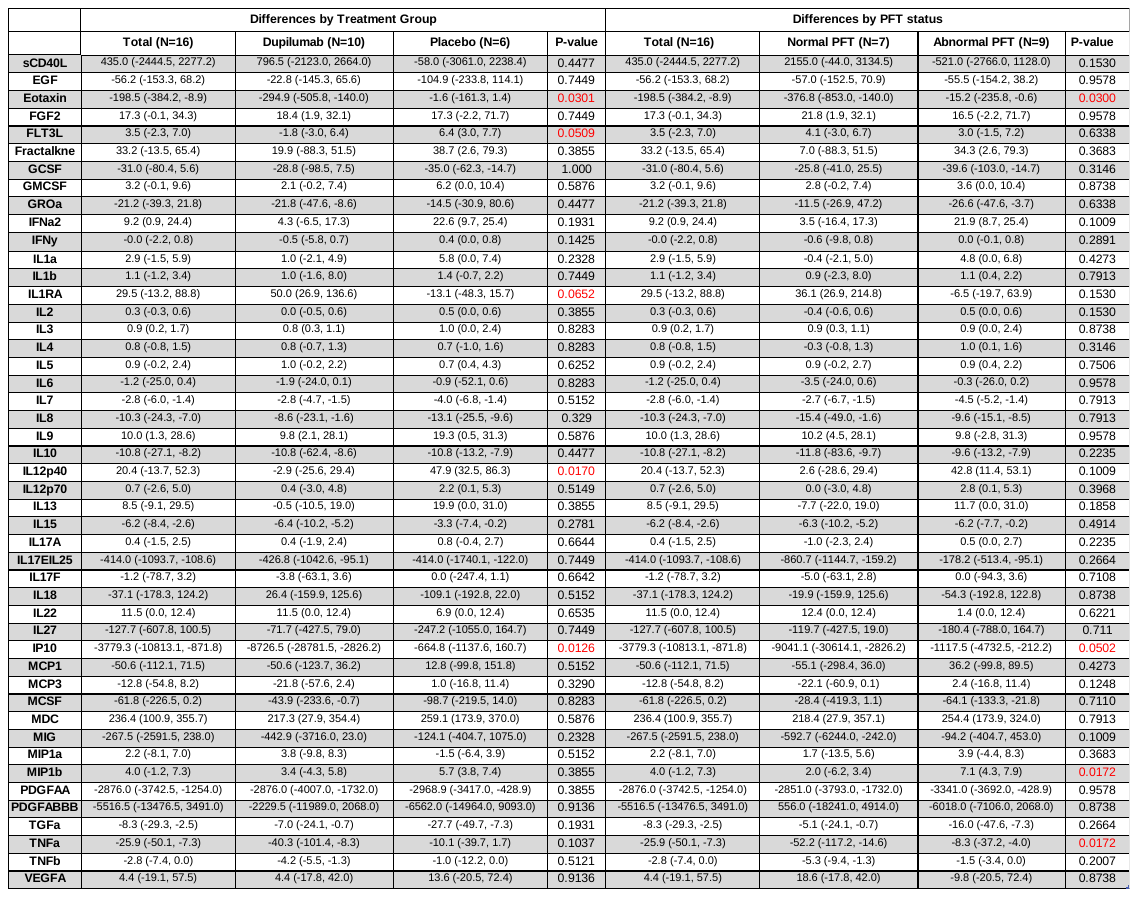

Table S5: Table showing median (interquartile range) (IQR) serum cytokine, chemokine or growth factor differences (column on far left) between day 365 and day 0 by treatment group (left side) and by PFT status (right side). P values are generated from Kruskal Wallis analyses. PFT= pulmonary function testing.
